# DNA methylation and brain dysmaturation in preterm infants

**DOI:** 10.1101/2021.04.08.21255064

**Authors:** Emily N. W. Wheater, Paola Galdi, Daniel L. McCartney, Manuel Blesa, Gemma Sullivan, David Q. Stoye, Gillian Lamb, Sarah Sparrow, Lee Murphy, Nicola Wrobel, Alan J. Quigley, Scott Semple, Michael J. Thrippleton, Joanna M. Wardlaw, Mark E. Bastin, Riccardo E. Marioni, Simon R. Cox, James P. Boardman

## Abstract

Preterm birth is associated with dysconnectivity of structural brain networks and is a leading cause of neurocognitive impairment in childhood. Variation in DNA methylation (DNAm) is associated with early exposure to extrauterine life but there has been little research exploring its relationship with brain development.

Using genome-wide DNA methylation data from saliva of 258 neonates, we investigated the impact of gestational age on the methylome and performed functional analysis to identify enriched gene sets from probes that contributed to differentially methylated probes (DMPs) or regions (DMRs). We tested the hypothesis that variation in DNAm could underpin the association between preterm birth and atypical brain development by linking DMPs with measures of white matter connectivity derived from diffusion MRI metrics: peak width of skeletonised mean diffusivity (PSMD), fractional anisotropy (PSFA) and neurite density index (PSNDI).

Gestational age at birth was associated with widespread differential methylation, with genome-wide significant associations observed for 8,870 CpG probes (*p*<3.6×10^−8^) and 1,767 differentially methylated regions. Functional analysis identified 14 enriched gene ontology terms pertaining to cell-cell contacts and cell-extracellular matrix contacts. Principal component analysis of probes with genome-wide significance revealed a first principal component (PC1) that explained 23.5% of variance in DNAm, and this was negatively associated with gestational age at birth. PC1 was associated with PSMD (β=0.349, *p*=8.37×10^−10^) and PSNDI (β=0.364, *p*=4.15×10^−5^), but not with PSFA (β=−0.035, *p*=0.510); these relationships mirrored the imaging metrics’ associations with gestational age at birth.

Gestational age at birth has a profound and widely distributed effect on the neonatal saliva methylome. Enriched gene ontology terms related to cell-cell contacts reveal pathways that could mediate the effect of early life environmental exposures on development. Finally, associations between differential DNAm and image markers of white matter tract microstructure suggest that variation in DNAm may provide a link between preterm birth and the dysconnectivity of developing brain networks that characterises atypical brain development in preterm infants.

## Introduction

Preterm birth, defined as birth at <37 weeks of gestation, affects around 11% of births worldwide^1^ and is a leading cause of neurodevelopmental and cognitive problems that extend across the life course. These include autism spectrum disorder, social difficulties, language impairment, ADHD, reduced IQ, educational underachievement, and psychiatric diagnoses.^2–9^

The neural phenotypes that underlie long-term functional impairment include diffuse white matter injury and subsequent dysmaturation of white matter and grey matter neuroaxonal structures, collectively termed the ‘encephalopathy of prematurity’.^10^ A consequence of the encephalopathy is generalised dysconnectivity of developing structural networks, which can be inferred from diffusion MRI (dMRI) and neurite orientation dispersion and density imaging (NODDI) during the neonatal period.^11–15^ Specifically, normal maturation is characterized by a reduction in mean diffusivity (MD) and increases in both fractional anisotropy (FA) and neurite density index (NDI) in white matter; but MD is increased and FA and NDI are decreased in preterm infants at term equivalent age, compared with healthy controls infants born at term.^16^ These changes reflect an increase in water content and a decrease in white matter organization in preterm infants. Peak width of skeletonised mean diffusivity (PSMD) is a method for histogram-based calculation of MD distribution across the entire white matter skeleton; it provides a measure of generalized white matter microstructure, is robust to scanner variation, and is predictive of cognition in later life.^17–19^ In previous work we extended the histogram model to neonatal data and included other dMRI and NODDI metrics. We found that PSMD and PSNDI are altered in preterm infants at term equivalent age, and that histogram-based measures have utility for investigating upstream determinants of dysmaturation such as systemic inflammation.^20,21^

The mechanisms that link the environmental stress of preterm birth with atypical brain development are uncertain. Variation in DNA methylation (DNAm) is a possible mechanism; DNAm is involved in the regulation of gene expression and cell fate during fetal brain development.^22^ Alterations in DNAm contribute to the pathogenesis of neurodevelopmental disorders (Rett syndrome,^23^ Immunodeficiency, Centromeric region instability, Facial anomalies [ICE] syndrome,^24^ and Angleman and Prader-Willi syndromes)^25^. There is growing evidence that differential DNAm can mediate the effect of environmental pressures on brain structure and function across the life course.^26,27^ The neonatal methylome is sensitive to prenatal factors such as maternal smoking,^28^ maternal body mass index,^29^ as well as birth weight.^30^ It is altered in association with co-morbidities of preterm birth,^26,31,32^ and there is some evidence for legacy differences in the methylome two decades after preterm birth.^33^ A meta-analysis investigating DNAm from umbilical cord blood identified widespread differential methylation associated with gestational age at birth across the gestational age range of 27-42 weeks, as measured on the Illumina 450k array.^34^ However, due to differences in the cellular composition of samples, epigenetic signatures observed in different tissues are likely to be distinct.^35^ The main cellular component of saliva, buccal epithelium, may be more representative of the brain than umbilical cord blood because of ectodermal origin.^36–38^

Here, our first aim was to determine whether gestational age at birth was associated with variation in the salivary methylome and to characterise the biological pathways implicated in the DNAm response to preterm birth. Our second aim was to investigate whether the DNAm signal of preterm birth explains variance in measures of white matter microstructure. We tested the hypotheses that gestational age at birth is associated with widespread differential methylation, and that DNAm contributes to variance in peak width skeletonised metrics of white matter connectivity, apparent during the neonatal period.

## Materials and methods

### Participants

Preterm and term born infants delivered at the Royal Infirmary of Edinburgh, UK were recruited to the Theirworld Edinburgh Birth Cohort, a longitudinal study designed to investigate the effect of preterm birth on brain development.^39^ Cohort exclusion criteria were major congenital malformations, chromosomal abnormalities, congenital infection, overt parenchymal lesions (cystic periventricular leukomalacia, hemorrhagic parenchymal infarction) or post-hemorrhagic ventricular dilatation. Ethical approval has been obtained from the National Research Ethics Service, South East Scotland Research Ethics Committee (11/55/0061, 13/SS/0143 and 16/SS/0154). Informed consent was obtained from a person with parental responsibility for each participant. The study was conducted according to the principles of the Declaration of Helsinki. DNAm data were available from 258 neonates, 214 of whom also had successful dMRI acquisition.

### DNA extraction and methylation measurement

Saliva obtained at term equivalent age was collected in Oragene OG-575 Assisted Collection kits, by DNA Genotek, and DNA extracted using prepIT.L2P reagent (DNA Genotek, Ontario, Canada). DNA was bisulfite converted and methylation levels were measured using Illumina HumanMethylationEPIC BeadChip (Illumina, San Diego, CA, USA) at the Edinburgh Clinical Research Facility (Edinburgh, UK). The arrays were imaged on the Illumina iScan or HiScan platform and genotypes were called automatically using GenomeStudio Analysis software version 2011.1 (Illumina). DNAm was analysed in two batches.

### DNA Methylation pre-processing

Raw intensity (.idat) files were read into R environment (version 3.4.4) using minfi. wateRmelon and minfi used for preprocessing, quality control and normalisation.^40,41^ The pfilter function in wateRmelon was used to exclude: samples with 1 % of sites with a detection p-value greater than 0.05; sites with beadcount <3 in 5% of samples; and sites with 1% of samples with detection p value >0.05. Cross hybridising probes and probes targeting single nucleotide polymorphisms with overall minor allele frequency ≥0.05 were also removed.^42^ Control probes were also removed. Samples were removed if there was a mismatch between predicted sex (minfi) and recorded sex (n = 3). Data was danet normalised which includes background correction and dye bias correction.^41^ Saliva contains different cells types, including buccal epithelial cells and leukocytes. Epithelial cell proportions were estimated with epigenetic dissection of intra-sample heterogeneity with the reduced partial correlation method implemented in the R package EpiDISH.^43^ Probes located on sex chromosomes were removed prior to analysis. Data from one of each twin pair was removed randomly (n=20).

### MRI acquisition

This study incorporates data from two phases of MRI acquisition. MRI was obtained at the same appointment as saliva sample collection for DNAm analysis. Structural and diffusion MRI were performed on 93 infants using a MAGNETOM Verio 3T clinical MRI scanner (Siemens Healthcare GmbH, Erlangen, Germany) and 12-channel phased-array head coil, which were used to acquire: dMRI employed a protocol consisting of 11 T2- and 64 diffusion-weighted (b = 750 s/mm^2^) single-shot spin-echo echo planar imaging (EPI) volumes acquired with 2 mm isotropic voxels (TE = 106 ms and TR = 7300 ms).

For the second phase (n=121), structural and diffusion MRI were performed on 121 infants using a MAGNETOM Prisma 3T clinical MRI scanner (Siemens Healthcare GmbH, Erlangen, Germany) and 16-channel phased-array pediatric head and neck coil were used to acquire dMRI in two separate acquisitions: the first consisted of 8 baseline volumes (b=0 s/mm^2^ [b0]) and 64 volumes with b=750s/mm^2^; the second consisted of 8 b0, 3 volumes with b=200 s/mm^2^, 6 volumes with b=500s/mm^2^ and 64 volumes with b = 2500 s/mm^2^. An optimal angular coverage for the sampling scheme was applied.^44^ In addition, an acquisition of 3 b0 volumes with an inverse phase encoding direction was performed. All dMRI volumes were acquired using single-shot spin-echo EPI with 2-fold simultaneous multislice and 2-fold in-plane parallel imaging acceleration and 2 mm isotropic voxels; all three diffusion acquisitions had the same parameters (TR/TE 3500/78.0ms). Images affected by motion artifact were re-acquired multiple times as required; dMRI acquisitions were repeated if signal loss was seen in 3 or more volumes.

Infants were fed and wrapped and allowed to sleep naturally in the scanner without sedation. Pulse oximetry, electrocardiography and temperature were monitored. Flexible earplugs and neonatal earmuffs (MiniMuffs, Natus) were used for acoustic protection. All scans were supervised by a doctor or nurse trained in neonatal resuscitation. Structural images were reported by an experienced pediatric radiologist (A.J.Q.)

### dMRI pre-processing

Diffusion images that were acquired on the MAGNETOM Verio scanner were denoised using a Marchenko-Pastur-PCA-based algorithm^45–47^; eddy current distortion and head movement were corrected using outlier replacement^48–50^; bias field inhomogeneity correction was performed by calculating the bias field of the mean b0 volume and applying the correction to all the volumes.^51^ FA and MD were calculated from the dMRI data.

The two dMRI acquisitions from the MAGNETOM Prisma scanner were first concatenated and then denoised using a Marchenko-Pastur-PCA-based algorithm^45–47^; eddy current, head movement and EPI geometric distortions were corrected using outlier replacement and slice-to-volume registration^48–50,52^; bias field inhomogeneity correction was performed by calculating the bias field of the mean b0 volume and applying the correction to all the other volumes.^51^ From the dMRI data we calculated the three eigenvalues and eigenvectors of the water diffusion tensor, and NODDI (Bingham distribution) parametric maps using cuDIMOT (intracellular volume fraction [NDI] and the overall orientation dispersion index [ODI_TOT_]).^12,13,53^ FA and MD were calculated using single-shell data to match the Verio scanner.

### Peak width of skeletonized water diffusion parameters

All the subjects were registered to the Edinburgh Neonatal Atlas (ENA50) using DTI-TK.^20^ The diffusion tensor derived maps of each subject (FA and MD) were calculated after registration; NDI was then propagated to the template space using the previously calculated transformations. The data was skeletonized using the ENA50 skeleton and then multiplied by a custom mask. Finally, the peak width of the histogram of values computed within the skeletonized maps was calculated as the difference between between the 95th and 5th percentiles.^17,20^

## Statistical analysis

### Epigenome-wide association analyses

Unless otherwise stated, analysis was completed in R version 3.4.4.^54^ Surrogate variable analysis of the data matrix was carried out, in order to adjust for potential confounders such as batch, using the sva function in the *sva* package in R.^55,56^ A fully adjusted model was specified prior to SVA to retain signal explained by biological variables of interest: CpG ~ Gestational age at birth + Gestational age at scan + Birthweight z score + Maternal smoking + Sex + Epithelial cells. SVA identified 17 significant surrogate variables which were subsequently included in the analysis.

EWAS was performed using the *limma* package in R.^57^ Beta values of each of 776025 CpG sites were regressed (as dependent variables) on gestational age at birth using linear regression. Covariates were added to adjust for sex, birthweight z score, gestational age at sample collection, maternal smoking, estimated epithelial cell proportions and surrogate variables. A significance threshold of 3.6×10^−8^ was selected, which represents genome-wide significance.^58^

### Differentially methylated region analysis

Differentially methylated regions were assessed using the dmrff function in the *dmrff* package in R.^59^ Here a differentially methylated region is a region containing two or above sites separated by ≤500 bp with EWAS analysis p ≤ 0.05 and methylation changes in a consistent direction. Following dmrff’s subregion selection step, DMRs with Bonferroni-adjusted p ≤ 0.05 were significant.

### Gene set testing

Gene set enrichment analysis was carried out using the GO and KEGG databases, and using the gometh function in *missMethyl* package, which controls for multiple probe bias.^60^ Sites that reached genome-wide significance in EWAS, and those that contributed to differentially methylated regions, were included in this analysis.

### Principal component analysis

Principal components analysis (PCA) was conducted on CpG probes that reached genome-wide significance, using the *prcomp* function in R. CpGs were pre-corrected for the effects of biological covariates and surrogate variables via linear regression. The scree plot was visually inspected in order to select a principal component (eigenvalue >1) to be carried forward for subsequent analysis.

### Linear regression between DNAm and PS metrics

Pearson’s correlation coefficient was used to assess the relationship between the first PC identified from PCA and gestational age at birth. This PC was used in linear regression models, as an independent variable, to test the associations between PSMD, PSNDI and PSFA with DNAm, conducted in R version 4.0.1.^54^ In models testing PSMD and PSFA, MRI scanner was included as a binary covariate as MRI data from both phases of data collection were included. PSNDI was only available for one phase of data collection and so it was not necessary to include scanner as a covariate. We report standardised regression coefficients and p-values.

### Data availability

The atlas with templates can be found at https://git.ecdf.ed.ac.uk/jbrl/ena and the code necessary to calculate histogram-based metrics is at https://git.ecdf.ed.ac.uk/jbrl/psmd. Requests for original image data will be considered through the BRAINS governance process: www.brainsimagebank.ac.uk.^61^ DNA methylation data will be deposited in NCBI’s Gene Expression Omnibus upon publication.

## Results

### Cohort

DNAm data were collected from 311 neonates. Twenty-nine did not meet DNAm preprocessing QC criteria and were excluded. One participant with a congenital abnormality was removed, as were three participants whose sex predicted from DNAm data did not match their recorded sex. This group of 278 neonates included 20 sets of twins. After random removal of one twin from each set there was no evidence of imbalance for birthweight (*t*=−0.157, *p*=0.88), or sex (Chi=0.417, *p*=0.52).

The study group consisted of 258 neonates: 155 participants were preterm and 103 were controls born at full term, see Table 1 for participant characteristics. Among the preterm infants, 38(25%) had bronchopulmonary dysplasia (defined as need for supplementary oxygen ≥36 weeks gestational age), 9(6%) developed necrotising enterocolitis requiring medical or surgical treatment, and 31(20%) had an episode of postnatal sepsis defined as either blood culture positivity with a pathogenic organism, or physician decision to treat for ≥5 days in the context of growth of coagulase negative staphylococcus from blood or a negative culture. Of the 258 participants with DNAm data, 214 also had MRI data.

**Table 1.** Participant characteristics

|  | **Preterm infants**<br>(n=155) | **Term infants**<br>(n=103) |
| --- | --- | --- |
| Gestational age at birth/weeks<br>(range) | 28.84<br>(23.28 – 34.84) | 39.7<br>(36.42 – 42.14) |
| Gestational age at scan/weeks<br>(range) | 40.56<br>(37.70-45.14) | 42.27<br>(39.84 – 47.14) |
| Birth weight/g<br>(range) | 1177<br>(500 – 2100) | 3482<br>(2346 – 4670) |
| Birth weight z-score<br>(range) | -0.19<br>(range -3.13 – 1.58) | 0.43<br>(range -2.30 – 2.96) |
| Sex: Female (%) | 75 (48) | 44 (43) |
| Maternal folate supplementation<br>in pregnancy (%) | 136 (88) | 86 (83) |
| Maternal age (years) | 31.1<br>(17–44) | 33.7<br>(19–48) |
| Maternal tobacco smoker in<br>pregnancy (%) | 29 (19) | 2 (2) |

### Widespread differential saliva DNAm in association with gestational age at birth

We conducted an epigenome-wide association study whereby CpG methylation at 776,025 sites was regressed on gestational age at birth, controlling for birthweight z-score, infant sex, gestational age at sample collection, maternal smoking, estimated epithelial cell proportion and surrogate variables. The genomic inflation factor was 1.72 (Supplementary Fig. 1). Differential methylation in relation to gestational age at birth was identified at 8,870 CpG sites at genome-wide significance (*p*<3.6×10^−8^), Fig. 1. Of these, 4,250 (47.9%) sites demonstrated a positive association with gestational age, while 4,620 (52.1%) were negatively associated. Following Bonferroni adjustment, 1,767 DMRs corresponding to 4,664 CpG sites were significant *p*<0.05. Of these, 11 had ten or more CpG sites contributing to the DMR. The largest DMR mapped to a genomic region that encodes two genes: *NNAT* and *BLCAP*. The 29 probes mapped to this region were all located within islands and positively associated with gestational age at birth, indicating that longer gestation corresponds to hypermethylation. Of the 10 most significant DMPs, 3 probes were localised to the *IRX4* gene, 1 probe to the *GAL3ST4* gene, and 1 to *LOXL4* (Table 2; Supplementary Fig. 2). The probes with the largest absolute magnitude effect size (top 5 hypermethylated and hypomethylated) mapped to the following genes: *IRX2, SMIM2, INTS1, HEATR2, ZBP1* and *UBXN11*, Table 3.

**Figure 1.**
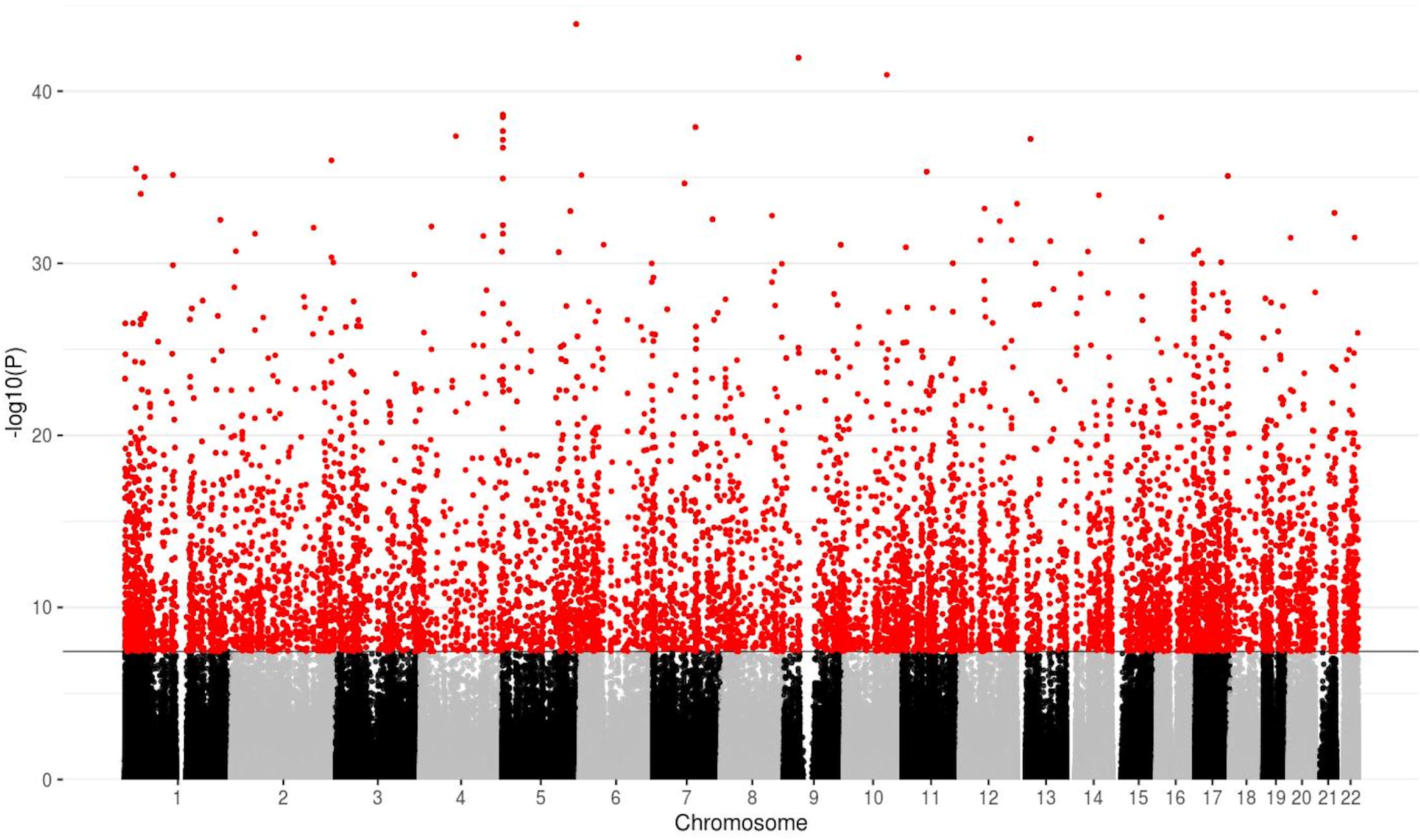
Manhattan plot for the association between gestational age at birth and DNA methylation, following adjustment for covariates and surrogate variables. The solid horizontal line shows the genome-wide significance level and red dots above this line represent probes that are significant at this threshold (*p*<3.6×10^−8^).

**Table 2.** Most significant probes associated with gestational age at birth.

| Probe | Chromosome | *p* value | Gene | Direction of effect | Coefficient* | Relation to island |
| --- | --- | --- | --- | --- | --- | --- |
| cg03558436 | 5 | $1.26 \times 10^{-44}$ | - | + | 0.01016 | Open Sea |
| cg04466438 | 9 | $1.13 \times 10^{-42}$ | - | + | 0.00755 | Open Sea |
| cg23701943 | 10 | $1.11 \times 10^{-41}$ | *LOXL4* | + | 0.01042 | Open Sea |
| cg18172877 | 5 | $2.31 \times 10^{-39}$ | *IRX4* | - | -0.00612 | Island |
| cg04180086 | 5 | $3.22 \times 10^{-39}$ | *IRX4* | - | -0.00736 | Island |
| cg22645539 | 7 | $1.22 \times 10^{-38}$ | *GAL3ST4* | - | -0.00815 | North Shelf |
| cg17774559 | 5 | $2.09 \times 10^{-38}$ | *IRX4* | - | -0.00635 | Island |
| cg17582074 | 4 | $4.12 \times 10^{-38}$ | - | + | 0.00525 | Open Sea |
| cg08915267 | 13 | $6.01 \times 10^{-38}$ | - | - | -0.00515 | North Shelf |
| cg04441405 | 5 | $6.60 \times 10^{-38}$ | - | - | -0.01102 | Island |

\*Coefficient corresponds to methylation change per week of gestation.

**Table 3.**
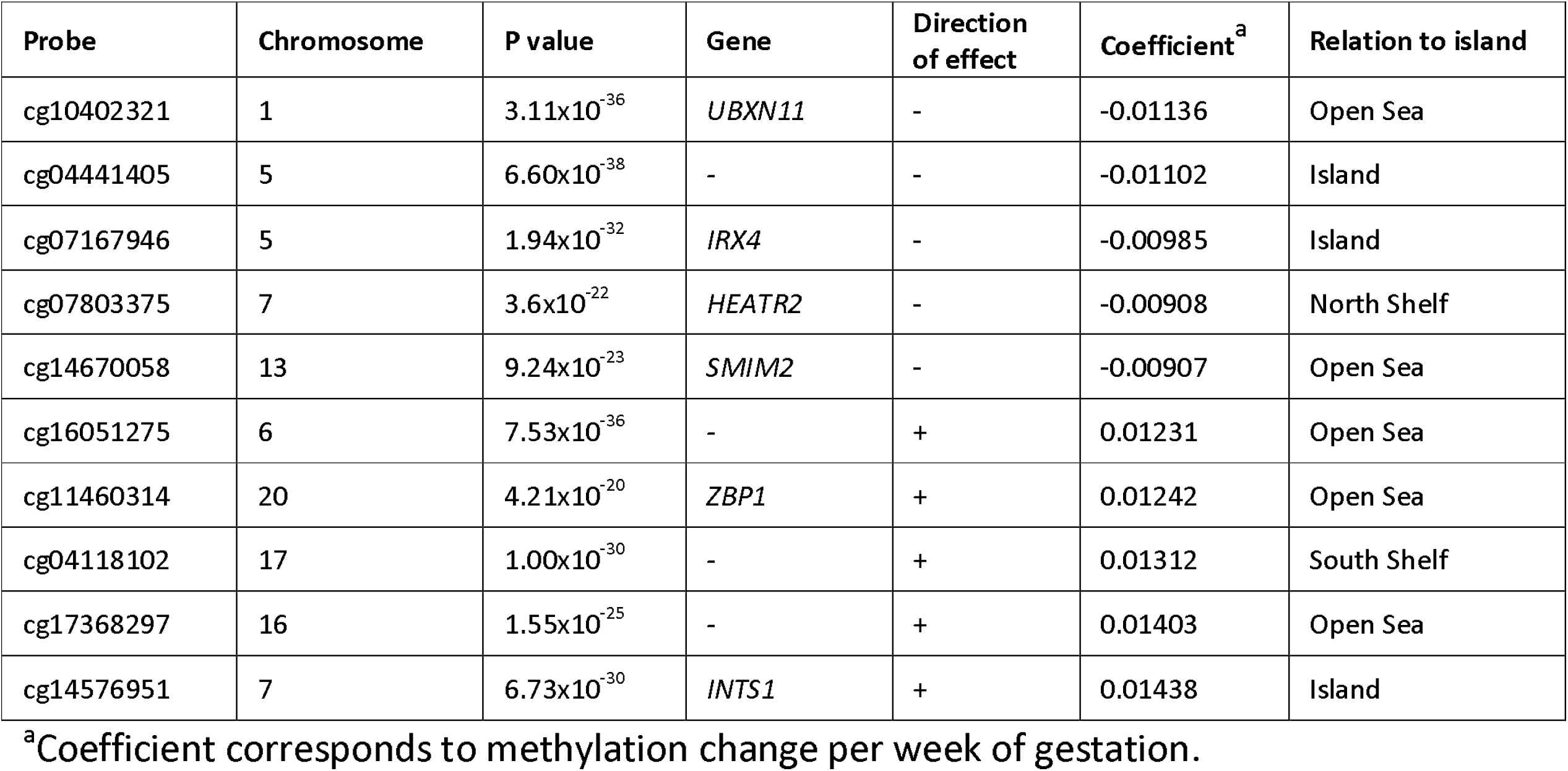
Probes with the largest absolute magnitude of association with gestational age at birth.

| Probe | Chromosome | P value | Gene | Direction of effect | Coefficient<sup>a</sup> | Relation to island |
| --- | --- | --- | --- | --- | --- | --- |
| cg10402321 | 1 | $3.11 \times 10^{-36}$ | *UBXN11* | - | -0.01136 | Open Sea |
| cg04441405 | 5 | $6.60 \times 10^{-38}$ | - | - | -0.01102 | Island |
| cg07167946 | 5 | $1.94 \times 10^{-32}$ | *IRX4* | - | -0.00985 | Island |
| cg07803375 | 7 | $3.6 \times 10^{-22}$ | *HEATR2* | - | -0.00908 | North Shelf |
| cg14670058 | 13 | $9.24 \times 10^{-23}$ | *SMIM2* | - | -0.00907 | Open Sea |
| cg16051275 | 6 | $7.53 \times 10^{-36}$ | - | + | 0.01231 | Open Sea |
| cg11460314 | 20 | $4.21 \times 10^{-20}$ | *ZBP1* | + | 0.01242 | Open Sea |
| cg04118102 | 17 | $1.00 \times 10^{-30}$ | - | + | 0.01312 | South Shelf |
| cg17368297 | 16 | $1.55 \times 10^{-25}$ | - | + | 0.01403 | Open Sea |
| cg14576951 | 7 | $6.73 \times 10^{-30}$ | *INTS1* | + | 0.01438 | Island |

<sup>a</sup>Coefficient corresponds to methylation change per week of gestation.

### Pathways implicated in functional testing

Based on the 8,870 sites that met the genome-wide significance threshold (*p*<3.6×10^−8^), no KEGG terms remained significant following FDR correction for multiple comparisons. Two Gene Ontology terms were enriched following FDR correction: one for anchoring (GO:0070161; q=0.0062) and one for adherens junction (GO:0005912; q=0.0062). In an analysis that incorporated all 11,752 distinct CpGs from both EWAS and DMR analysis, 14 GO terms were enriched, Table 4.

**Table 4.**
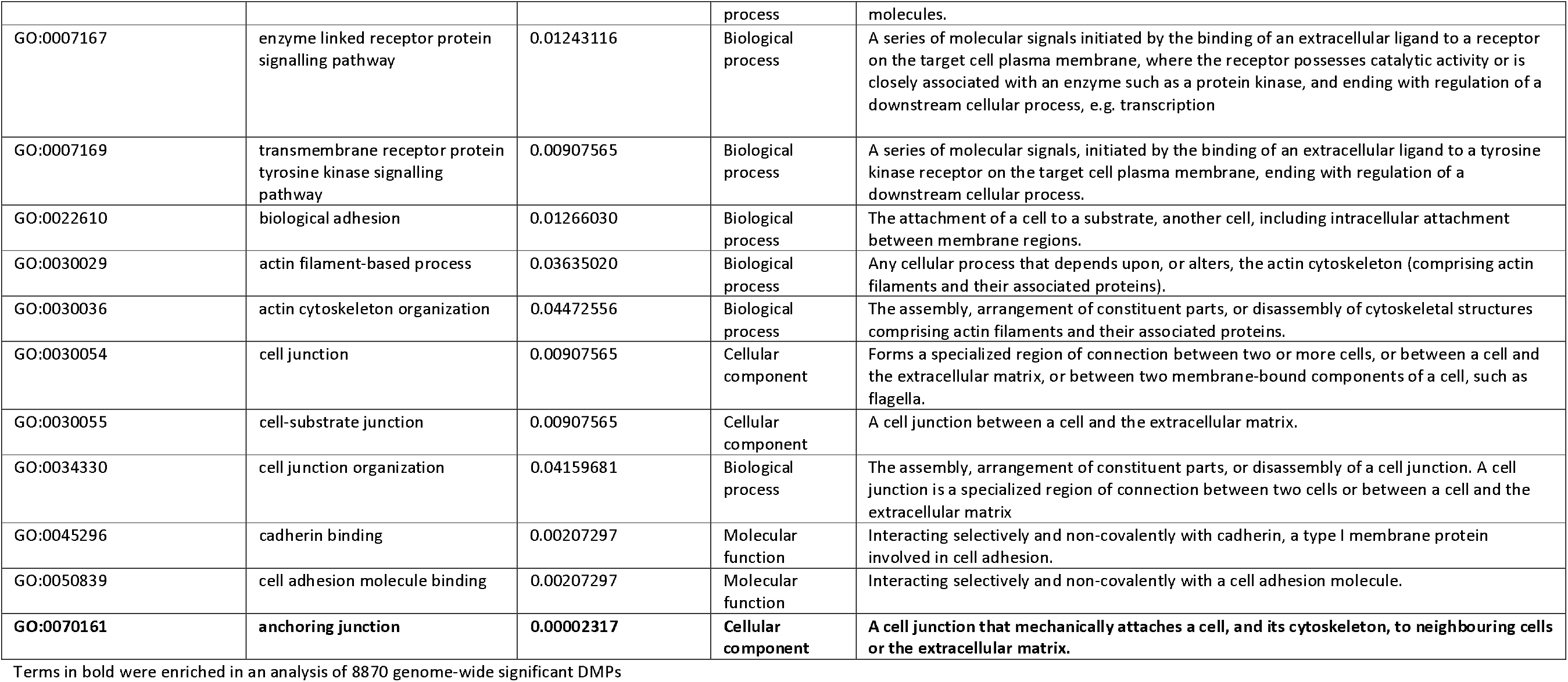
Gene ontology (GO) terms that were significantly enriched in an analysis of all probes that contributed to DMPs and DMRs.

|  |  |  | process | molecules. |
| --- | --- | --- | --- | --- |
| GO:0007167 | enzyme linked receptor protein signalling pathway | 0.01243116 | Biological process | A series of molecular signals initiated by the binding of an extracellular ligand to a receptor on the target cell plasma membrane, where the receptor possesses catalytic activity or is closely associated with an enzyme such as a protein kinase, and ending with regulation of a downstream cellular process, e.g. transcription |
| GO:0007169 | transmembrane receptor protein tyrosine kinase signalling pathway | 0.00907565 | Biological process | A series of molecular signals, initiated by the binding of an extracellular ligand to a tyrosine kinase receptor on the target cell plasma membrane, ending with regulation of a downstream cellular process. |
| GO:0022610 | biological adhesion | 0.01266030 | Biological process | The attachment of a cell to a substrate, another cell, including intracellular attachment between membrane regions. |
| GO:0030029 | actin filament-based process | 0.03635020 | Biological process | Any cellular process that depends upon, or alters, the actin cytoskeleton (comprising actin filaments and their associated proteins). |
| GO:0030036 | actin cytoskeleton organization | 0.04472556 | Biological process | The assembly, arrangement of constituent parts, or disassembly of cytoskeletal structures comprising actin filaments and their associated proteins. |
| GO:0030054 | cell junction | 0.00907565 | Cellular component | Forms a specialized region of connection between two or more cells, or between a cell and the extracellular matrix, or between two membrane-bound components of a cell, such as flagella. |
| GO:0030055 | cell-substrate junction | 0.00907565 | Cellular component | A cell junction between a cell and the extracellular matrix. |
| GO:0034330 | cell junction organization | 0.04159681 | Biological process | The assembly, arrangement of constituent parts, or disassembly of a cell junction. A cell junction is a specialized region of connection between two cells or between a cell and the extracellular matrix |
| GO:0045296 | cadherin binding | 0.00207297 | Molecular function | Interacting selectively and non-covalently with cadherin, a type I membrane protein involved in cell adhesion. |
| GO:0050839 | cell adhesion molecule binding | 0.00207297 | Molecular function | Interacting selectively and non-covalently with a cell adhesion molecule. |
| **GO:0070161** | **anchoring junction** | **0.00002317** | **Cellular component** | **A cell junction that mechanically attaches a cell, and its cytoskeleton, to neighbouring cells or the extracellular matrix.** |

Terms in bold were enriched in an analysis of 8870 genome-wide significant DMPs

### Gestational age at birth is associated with metrics of white matter microstructure in neonates

DNAm and PSMD and PSFA were both available for 214 infants (Supplementary Table 1), and DNAm and PSNDI were available for the 121 infants from phase 2 (Supplementary Table 2). Gestational age at birth was significantly associated with PSMD (β=−0.602, *p*<2×10^−16^) and PSNDI (β=−0.594, *p*=2.17×10^−9^), but not with PSFA (β=−0.005 *p*=0.933), Fig. 2A and 2B; Table 5.

**Figure 2.**
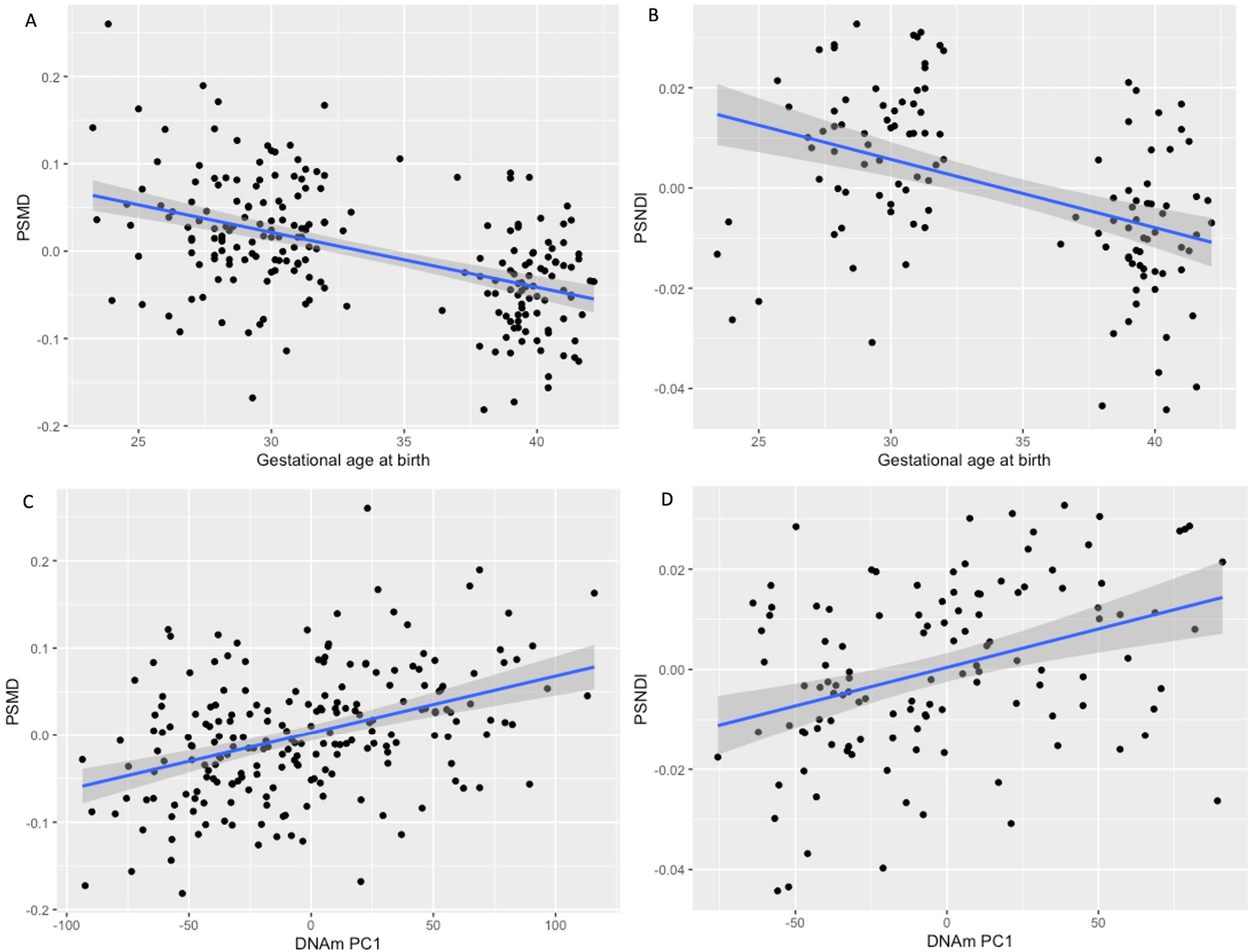
Scatter plots with 95% confidence intervals, showing the relationships between gestational age at birth and DNAm with peak width skeletonised mean diffusivity and neurite density index. The associations between gestational age and PSMD and PSNDI are shown in A and B, respectively. The relationships between DNAm PC1 and PSMD and PSNDI are shown in C and D, respectively. PS metrics are residualised for gestational age at scan; PSMD is additionally residualised for scanner variable.

**Table 5.** Associations between global metrics of white matter microstructure, DNAm first principal component (left) and gestational age (right)

| PS metric | Metric ~ PC1 DNAm + Gestational age at scan + scanner variable<sup>a</sup> |  | Metric ~ Gestational age at birth + Gestational age at scan + scanner variable<sup>a</sup> |  |
| --- | --- | --- | --- | --- |
| | $\beta$ | $p$ | $\beta$ | $p$ |
| PSFA | -0.035 | 0.510 | -0.005 | 0.933 |
| PSMD | 0.349 | **8.37x10<sup>-10</sup>** | **-0.602** | **< 2x10<sup>-16</sup>** |
| PSNDI | 0.364 | **4.15x10<sup>-5</sup>** | **-0.594** | **2.17x10<sup>-9</sup>** |

<sup>a</sup>scanner variable was included in the model for PSFA and PSMD but not PSNDI because NODDI imaging was carried out for a subset using a single MRI scanner (n=121). Bold print signifies significant associations.

### Differential DNAm is associated with white matter microstructure

The first unrotated PC derived from the 8,870 genome-wide significant CpGs accounted for 23.5% of the variance, the second PC accounted for 2.5%, Fig. 3A. There was no evidence of batch effects in the PCs (Supplementary Fig. 3). PC1 was significantly correlated with gestational age at birth (r=−0.622; *p*<2.2×10^−16^), Fig. 3B. PC1 was also positively associated with PSNDI (β=0.364, *p*=4.15×10^−5^), and in models adjusted for scanner it was positively associated with PSMD (β=0.349, *p*=8.37×10^−10^) but not PSFA (β=−0.035, *p*=0.510), Table 5. All models were adjusted for gestational age at scan.

**Figure 3.**
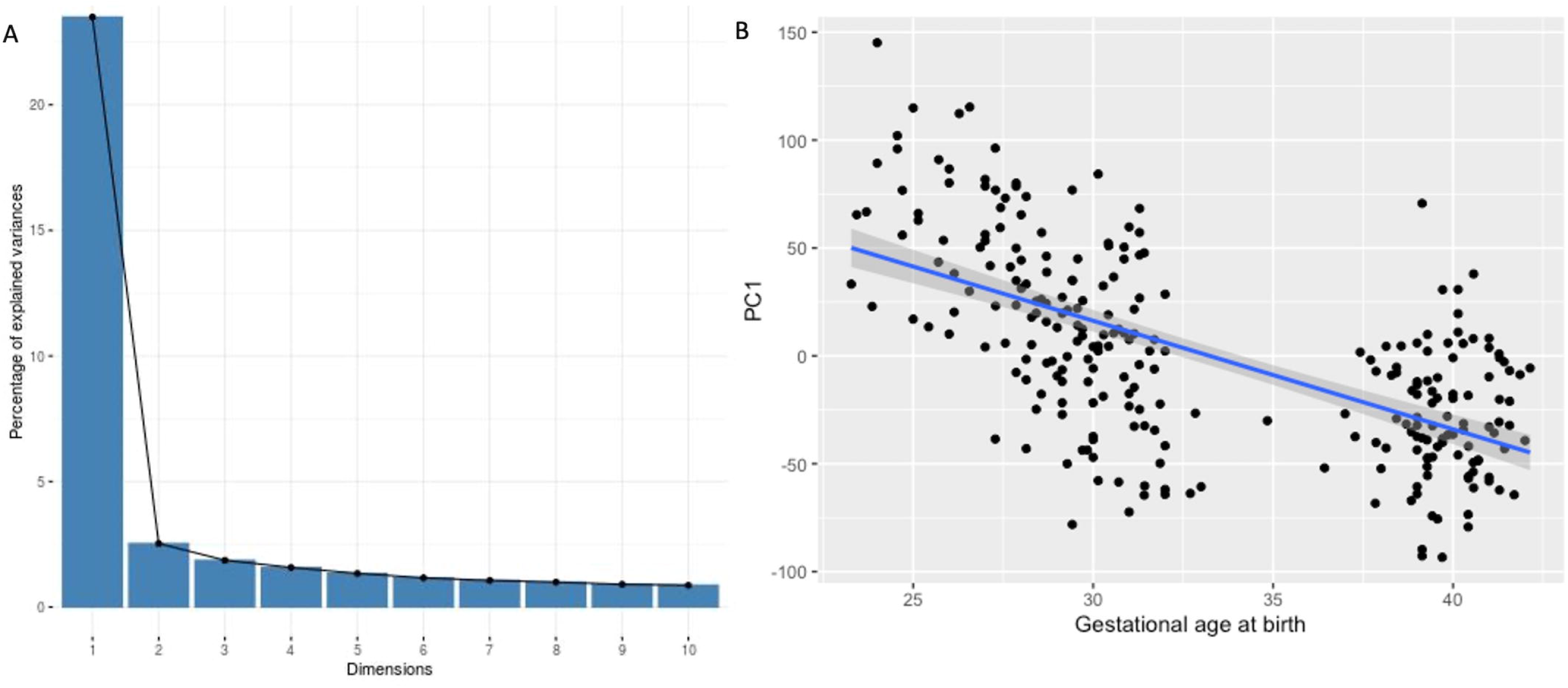
Variation in DNAm probes selected by EWAS captured by principal component, and the relationship with gestational age. Fig 3A, scree plot showing sharp elbow after the first PC, accounting for 23.5% of variance. Fig 3B, scatter plot showing relationship between gestational age at birth and PC1 (r=-0.622) with 95% confidence intervals.

## Discussion

By studying a unique database of DNA linked to brain imaging in a population of preterm infants and healthy controls, we have identified extensive differential methylation in association with gestational age at birth and revealed an association between the principal axis of methylation and brain dysmaturation within the same sample. Differentially methylated regions and probes were distributed widely across the genome, indicating that gestation duration has a global effect on DNA methylation. Gene enrichment analysis of changes associated with gestational age identified gene sets pertaining to cell contacts and cytoskeleton. A single principal component that explained 23.5% of the variance in differential DNAm linked to preterm birth was closely associated with markers of generalised dysconnectivity across the white matter skeleton.

The data are consistent with studies that have reported associations between length of gestation and genome-wide variation in DNAm within fetal brain ^22^ and umbilical cord blood ^34,62^; and widely distributed variation is reported in association with postmenstrual age at sampling of preterm infants (a proxy for GA at birth).^31^ The signature we identified in saliva sampled at term equivalent age included 233 probes that were previously shown to be differentially methylated in association with GA in a meta-analysis of umbilical cord blood samples that reported 8,899 gestation-dependent CpGs.^34^ The limited overlap could be explained by differences in cellular composition of assessed tissues, different array types used to measure DNAm, or due to the time of sampling. Cord blood is collected at birth and so methylation changes at this time reflect fetal maturity and/or prenatal experience, whereas the methylation signature at term equivalent age reflects the allostatic load of early postnatal experiences as well as the prenatal environment. We chose to sample at term equivalent age because postnatal co-morbidities of preterm birth and NICU care practices such as painful stress exposures alter DNAm profiles, and because cumulative DNAm variations over this time period may link exposure to behavioural outcome in preterm infants.^32,63,64^

Functional analyses of DMPs identified two enriched gene ontology terms, for adherens and anchoring junctions. When distinct probes that contributed to both DMPs and DMRs were combined, gene ontology analysis identified an additional 14 terms related to cell-cell adhesion, cell adhesions with the extracellular matrix, and signalling from the extracellular membrane; 12 of these were also identified in the meta-analysis of gestational age effects on DNAm obtained at birth from umbilical cord blood.^34^ The most significant DMR mapped to a site encoding two genes including *NNAT*, which encodes the neural fate initiator neuronatin, the expression of which decreases throughout development ^65^; there was a positive association with increasing gestational age at birth. Hypomethylation of *NNAT* is associated with a corresponding increase in expression of neuronatin.^66^ Furthermore, the candidacy of *NNAT* as a gene of interest whose expression may be modified by perinatal exposures is supported by previous EWAS of gestational age, maternal body mass index, maternal smoking, and schizophrenia.^22,29,62,67–70^

Probes that demonstrated the largest magnitude of effect in association with gestational age mapped to genes previously associated with gestational age or maternal risk factors in EWAS.^70^ Hypermethylated genes include: *ZBP1*, which was identified in EWAS investigating gestational age and hypertensive disorders of pregnancy ^62,69,71^; *INTS1*, which has been identified in EWAS of gestational age, hypertensive disorders of pregnancy, maternal body-mass-index, birth weight and breastfeeding duration.^22,30,62,67,71,72^ Hypomethylated genes included: *UBXN11*, which was identified in studies of gestational age ^22,62^; and *IRX4*. Three of the ten most significant DMPs mapped to the *IRX4* gene, all of which displayed a negative association with gestational age at birth. *IRX4* is associated with cardiac development in vertebrates, including humans.^73^ Its homologues have been implicated in retinal axon guidance in zebrafish, and neural patterning in *Xenopus* ^74,75^, and it has been identified in previous EWAS of hypertensive disorders of pregnancy ^71^ and prenatal maternal stress.^76^

The novel pathways and genes implicated by EWAS studies of gestational age could provide a strong empirical basis for the selection of genes in targeted analyses in association with neuroimaging.^27^ For example, one of the genes identified in our EWAS has been implicated in neurodevelopmental disorders; biallelic mutations in *INTS1* have been associated with a rare neurodevelopmental syndrome characterised by intellectual disability.^77–79^

We used metrics of generalised white matter connectivity to assess relationships between DNAm and brain development because generalised dysconnectivity of structural networks is a hallmark of the preterm brain dysmaturation.^10,14,80^ PSNDI and PSMD were strongly associated with the first principal component of GA-dependent variation in DNAm, but PSFA was not. This suggests that variations in DNAm could contribute to the higher variability in water content and intra-axonal volume that characterise preterm brain dysmaturation.^20^ We have previously reported that differential DNAm is associated with FA of the genu of the corpus callosum and tract shape of the right corticospinal tract ^26^; it is most likely that we did not observe an association between neither GA nor DNAm with PSFA because this metric is subject to histogram shift ^20^, meaning that although there are groupwise differences in FA values across the skeleton, the spread of values is the same. Associations between GA at birth and both DNAm and image markers of dysconnectivity, and between DNAm and image features, suggest that differential DNAm contributes, in part, to the relationship between GA and brain network dysconnectivity in preterm infants.

Strengths of this study are that we studied a population of preterm and control infants across the gestational age range of 23 to 42 weeks, who were uniquely phenotyped with DNAm and dMRI. We sampled after the period of NICU care in order to capture the allostatic load of preterm birth. We measured DNAm from neonatal saliva samples, which has consistency with brain DNAm patterns and is non-invasive. The Illumina EPIC platform provided extensive coverage of the methylome (850,000 sites) and we controlled for cell composition. Finally, we used an image phenotype that is robust to scanner variation.^17^ There are some limitations. First, control for cell composition was based on estimation of cell proportions rather than measurement, so we cannot rule out the possibility that some of the signal identified was related to variation in cell composition. Second, mediation analysis to assess causation was not possible because the association between the DNAm PC1 and PS metrics might result from the DNAm PC being derived from CpG sites that are associated with gestational age, so variance attributable to the mediating variable cannot be assumed. This could be addressed by out-of-sample validation, which will require other neonatal cohorts with both methylome and dMRI data. There was also some evidence of inflation of test statistics based on the genomic inflation factor. However, genomic inflation factor is thought to provide an overestimate of inflation and corrections based on it may be overly conservative.^81^ The value of the genomic inflation factor was also similar to those previously reported in neonatal cohorts.^34^ In addition, visual inspection of our results via Manhattan plots suggests that our finding, of widespread differences in DNAm in relation to gestational age at birth, are in line with previous studies that have investigated this in cord blood and in fetal brain tissue.^22,34^

In conclusion, gestational age at birth has a profound and widely distributed impact on the neonatal saliva methylome at term equivalent age, which reflects the allostatic load of preterm birth itself and postnatal exposures during neonatal intensive care. Gene ontology terms related to cell-cell contacts were enriched, indicating that cell contacts and organisation are implicated in the phenotype. Associations between DNAm and PSMD and PSNDI suggest that variations in DNAm could contribute to white matter dysconnectivity commonly seen in preterm infants, and this analysis identified several genes and gene regions that could provide further insight into the molecular mechanisms by which early exposure to extrauterine life influences neurodevelopment.

## Supporting information

STROBE_checklist_Wheater_2021

SupplementaryMaterial_Wheater_2021

## Data Availability

The atlas with templates can be found at https://git.ecdf.ed.ac.uk/jbrl/ena and the code necessary to calculate histogram-based metrics is at https://git.ecdf.ed.ac.uk/jbrl/psmd. Requests for original image data will be considered through the BRAINS governance process: www.brainsimagebank.ac.uk. DNA methylation data will be deposited in NCBI’s

Gene Expression Omnibus upon publication.

https://git.ecdf.ed.ac.uk/jbrl/ena

https://git.ecdf.ed.ac.uk/jbrl/psmd

http://www.brainsimagebank.ac.uk/

## Acknowledgements

Some of the participants were scanned in the University of Edinburgh Imaging Research MRI Facility at the Royal Infirmary of Edinburgh which was established with funding from The Wellcome Trust, Dunhill Medical Trust, Edinburgh and Lothians Research Foundation, Theirworld, The Muir Maxwell Trust, and other sources. We thank Thorsten Feiweier at Siemens Healthcare for collaborating with dMRI acquisitions (Works-in-Progress Package for Advanced EPI Diffusion Imaging).

We are grateful to the families who consented to take part in the study and to all the University’s imaging research staff for providing the infant scanning.

## Abbreviations

ADHD: attention deficit hyperactivity disorder
FA: fractional anisotropy
IQ: intelligence quotient
MD: mean diffusivity
NODDI: neurite orientation dispersion and density imaging
NDI: neurite density index
PS: peak width skeletonised
PCA: principal component analysis
SVA: surrogate variable analysis.

---

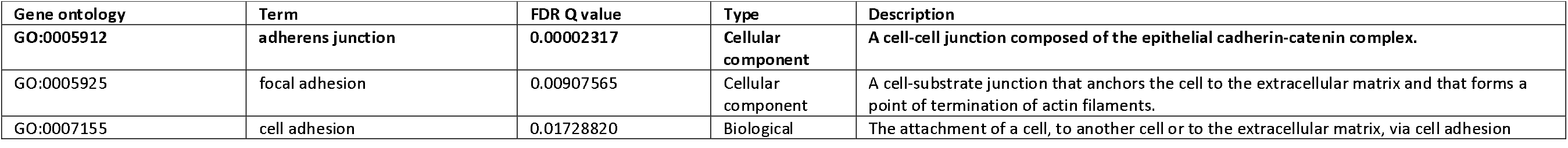

| Gene ontology | Term | FDR Q value | Type | Description |
| --- | --- | --- | --- | --- |
| GO:0005912 | adherens junction | 0.00002317 | Cellular component | A cell-cell junction composed of the epithelial cadherin-catenin complex. |
| GO:0005925 | focal adhesion | 0.00907565 | Cellular component | A cell-substrate junction that anchors the cell to the extracellular matrix and that forms a point of termination of actin filaments. |
| GO:0007155 | cell adhesion | 0.01728820 | Biological | The attachment of a cell, to another cell or to the extracellular matrix, via cell adhesion |

