## SupplementaryMaterial_Wheater_2021 for "DNA methylation and brain dysmaturation in preterm infants"

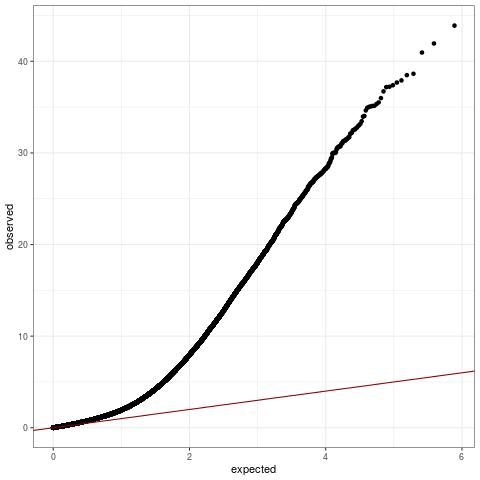

**Supplementary Figure 1. Quantile-Quantile (QQ) plot for EWAS results. Genomic inflation factor: 1.72.**

**Supplementary Figure 2**. Scatter plots showing the relationship between the beta values of top ten most significant CpG probes and gestational age at birth in weeks, with 95% confidence intervals.

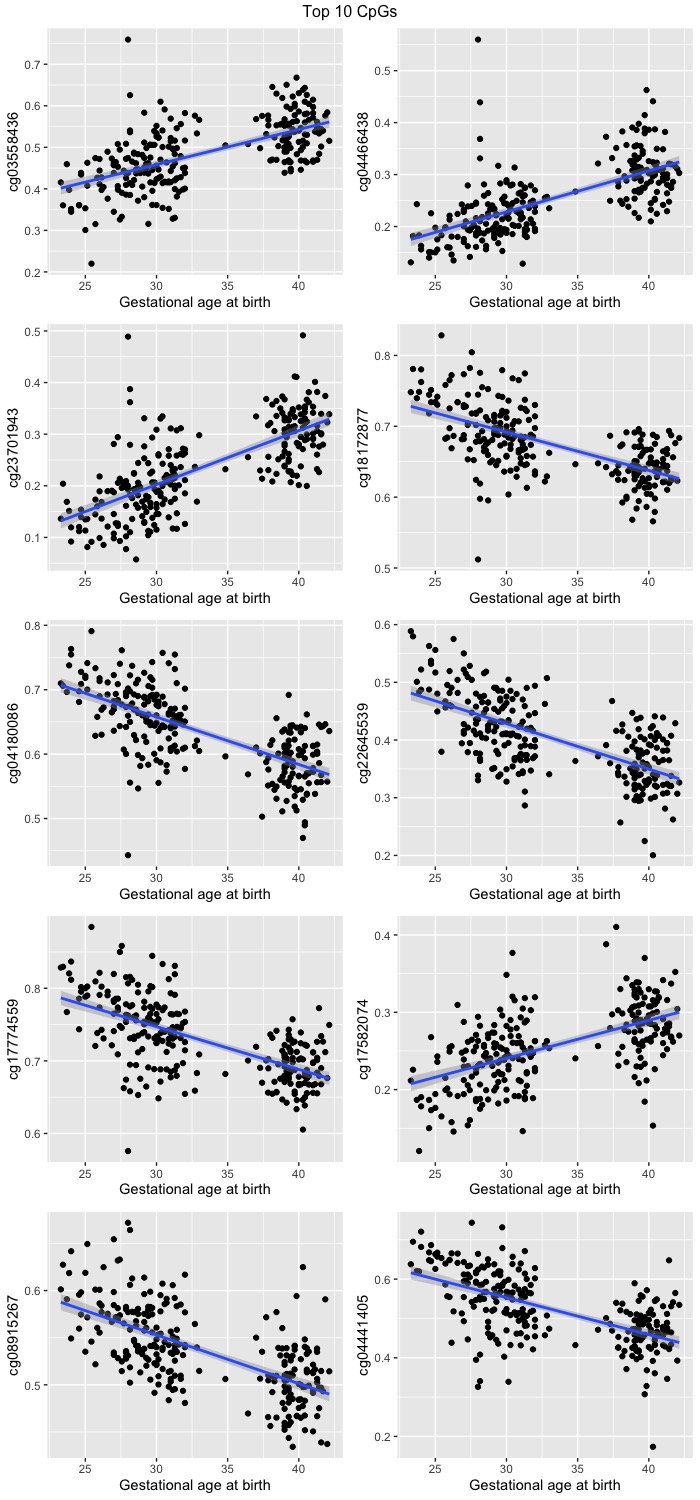

**Supplementary Figure 3.** Batch effects have been successfully removed from the residualised DNAm data based on the top 2 PCs derived from 8,870 CpG probes that reached genome-wide significance.

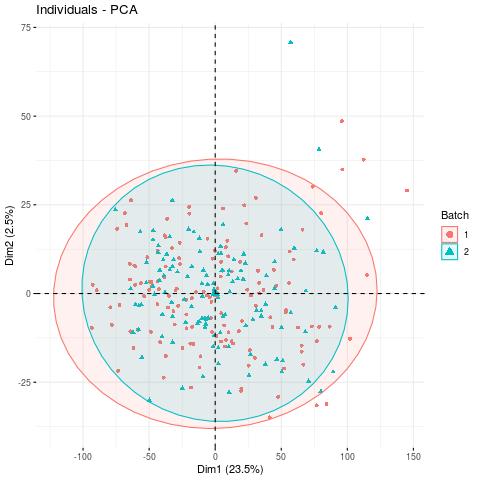

**Supplementary Table 1.** Demographics of participants with both PS (FA and MD) metrics and DNAm data

|  | **Preterm Infants**  (n=127) | **Term Infants**  (n=87) |
| --- | --- | --- |
| Gestational age at birth/weeks (range) | 29.14  (23.28 – 34.86) | 39.74  (36.43 – 42.14) |
| Gestational age at scan/weeks (range) | 40.44  (37.70-44.29) | 42.25  (39.84- 47.14) |
| Birth weight/g  (range) | 1184  (500 – 2100) | 3489  (2410 -4670) |
| Birth weight z-score  (range) | -0.2476  (-3.1324 – 1.5809) | 0.4521  (-2.2952 – 2.9620) |
| Maternal folate supplementation in pregnancy (%) | 110  (87) | 76  (87) |
| Sex: Female (%) | 63 (50) | 39 (45) |
| Maternal age (years) | 31.0  (17-44) | 33.9  (23 – 45) |
| Maternal tobacco smoker in pregnancy (%) | 24 (19) | 0 (0) |

**Supplementary Table 2.** Demographics of participants with both PSNDI metric and DNAm data

|  | **Preterm Infants**  (n=64) | **Term Infants**  (n=57) |
| --- | --- | --- |
| Gestational age at birth/weeks (range) | 29.28  (23.43 – 32.00) | 39.71  (36.43 – 42.14) |
| Gestational age at scan/weeks (range) | 40.44  (38.29-44.29) | 42.25  (40.00- 47.14) |
| Birth weight/g  (range) | 1266  (500 – 2100) | 3502  (2410 -4560) |
| Birth weight z-score  (range) | -0.1083  (-3.1324 – 1.5809) | 0.4996  (-2.2952 – 2.5703) |
| Maternal folate supplementation in pregnancy (%) | 50  (78) | 47  (82) |
| Sex: Female (%) | 25 (39) | 25 (44) |
| Maternal age (years) | 31.2  (21-44) | 34.2  (23-44) |
| Maternal tobacco smoker in pregnancy (%) | 11 (17) | 0 (0) |
